# Early CPAP reduced mortality in covid-19 patients. Audit results from Wrightington, Wigan and Leigh Teaching Hospitals NHS Foundation Trust

**DOI:** 10.1101/2020.05.28.20116152

**Authors:** Abdul Ashish, Alison Unsworth, Jane Martindale, Luigi Sedda, Ramachandran Sundar, Martin Farrier

## Abstract

COVID-19 infection typically causes pneumonia with bilateral changes on Chest radiograph. There is significant hypoxia and use of oxygen for patients admitted to hospital is standard. The use of Continuous Positive Airway Pressure (CPAP) in patients with COVID-19 has now become established as a common clinical practice based on recent experience. It is given as part of “best endeavours” treatment in the absence of sufficient evidence to guide best practice. The use of CPAP as a step up in clinical care is now common but has a poor evidence base.

Using routinely collected data, the use of CPAP as a supportive non-invasive ventilatory treatment is described in 35 patients with COVID infection. Patients given early CPAP and in particular within 48 hours of admission, are shown to have a better outcome (a significant probability of lower mortality) than patients who received late CPAP (more than 48 hours after admission).

Although the analysis is affected by a small sample size, the results have shown good evidence that supports the early use of CPAP in patients with COVID-19 infection.

## Background

COVID-19 was declared a Pandemic by the World Health Organisation on 12^th^ March 2020. This led to a major outbreak in the United Kingdom. Knowledge about treatment was limited as the pandemic progressed (Nicola et al. 2020), while the first general guidelines were provided by NICE on 20^th^ of March (https://www.nice.org.uk/guidance/ng159).

Wrightington Wigan and Leigh Teaching Hospitals NHS Trust (hereafter simply the Trust) is a medium sized organisation caring for a population of 330,000 people which admitted the first patients with COVID-19 on 14^th^ March 2020. By 18^th^ May 2020 there had been 504 patients admitted to the Trust with COVID-19 as diagnosed using antigen testing from nasal and oral swabs.

As patients began to be admitted, there was very little evidence on the effectiveness of CPAP in Acute Respiratory Failure (ARF) (Williams et al. 2013; Bakke et al. 2014; Faria et al. 2015). There was however some international evidence and recommendations beginning to emerge, that CPAP treatment and high flow nasal oxygen was promising (Rochwerg, 2019, Zhonghua, 2020). It was also suggested that early intubation of a patient with known or suspected COVID-19 and respiratory distress could have been too early as patients may have improved on CPAP or Non Invasive Ventilation (Arulkumaran, 2020).

Within the Trust, CPAP was used very little in the early weeks of the COVID outbreak, but national and international experience indicated that CPAP was useful for patients. Therefore a decision was taken to use CPAP on a much larger scale, which in itself was a huge challenge for the Trust. On a logistical basis, increasing the number of CPAP machines and identification of designated bays on wards was the primary concern. Larger Trusts provided CPAP use on their ICU/HDUs due to the complexity of these patients’ clinical picture and care needs. As a ‘smaller’ Trust with fewer ICU/HDU beds, this required our nursing staff to quickly upskill on the use of CPAP on medical ward with adaptable protocols and risk assessments. This was overseen by the Respiratory team and supported by the intensive care “outreach team”. The Trust outreach teams were pivotal at this stage sharing their expertise to educate, support and guide throughout this process.

The CPAP machines chosen were Resmed Stellar devices. These are designed to deliver non-invasive ventilation for inpatients admitted with Type 2 respiratory failure and chronic obstructive airway disease.. The machines and the facemasks were adapted for use in COVID-19 patients to minimise aerosol generation. Oxygen was delivered at 5 to 15 litres / min through via a port in the facemask. This delivers an estimated 50 – 70% FiO2.

Patients were selected for CPAP therapy by Respiratory team according to the unit protocol at the time. The protocols were adapted continuously as clinical expertise and confidence developed. CPAP initiation in these patients developed over the course of the first few weeks and an earlier initiation of CPAP was trialled based on emerging experience.

Early CPAP therapy was considered to be given if a patient was started on CPAP treatment within 48 hours of admission and late CPAP was defined as being started after 48 hours. It was possible to capture timing of the start of CPAP due mandatory recording as part of the nursing observations in the Clinical Notes in the Electronic Patient Records from the start of the outbreak.

Given the absence (at the moment this manuscript was prepared – end of May 2020) of any quantitative assessment on the effect of the use of CPAP in patient’s mortality, and given the increasing expertise in managing patients with CPAP by the personnel of the Trust, the decision was made to analyse the effectiveness of this intervention by registering to complete a clinical audit (DB ref 4639) drawing data from the Electronic Patient Record System. It was possible to track all patients who had received treatment with CPAP therapy and at what stage of their admission.

This short manuscript describes the data collection and statistical methods employed to evaluate if early introduction of CPAP as opposed to standard care recommendations, could lower the morality rate from COVID-19 and prevent the need for ventilation.

## Data and Methods

### Patient selection

All patients admitted with COVID-19 infection who were started on CPAP between 6^th^ March 2020 and 6^th^ May 2020 were included. There were a total of 35 patients, 12 females and 13 males. The median age was 58 years with minimum of 34 and maximum 81 years.

The information available for each patient was: sex, age, outcome (died or survived), oxygen (l/min), and time when the CPAP was used (in days). Age was transformed in a binary variable (A50) with value of 1 for patients older than 50 years old and 0 for patients younger than 50 years old (this threshold was based on the goodness of fit of the mixed model applied below). From time of CPAP was created an additional variable, early CPAP, with values of 1 when CPAP was performed within the first two days, and the value of 0 otherwise.

### Statistical analysis

For this analysis we have employed a Binomial (logit) generalised mixed model (Pinheiro and Bates, 2000), with patient outcome as response, and for the fixed effect the time of CPAP (continuous), squared Oxygen (continuous) and early CPAP (binary). For the random effect A50 within sex was chosen.

The describe model components allowed for the model with lowest BIC (Delattre et al. 2012).

## Results

Table 1 shows the results for the fixed effect of the Binomial (logit) generalised linear mixed model.

**Table 1.** Results for the fixed effect of the Binomial (logit) generalised linear mixed model. Odd ratios and 95 confidence interval are reported in the last three columns.

| Predictor | Coefficient | P-value | OR | Lower limit | Upper limit |
| --- | --- | --- | --- | --- | --- |
| CPAP time | -0.133 | 0.041 | 0.875 | 0.769 | 0.994 |
| Oxygen<sup>2</sup> | 0.013 | 0.009 | 1.014 | 1.003 | 1.024 |
| CPAP early | -2.751 | 0.011 | 0.064 | 0.007 | 0.536 |

Both CPAP time – e.g. when CPAP was given to the patient (in number of days after hospitalization), and early CPAP (CPAP within two days from hospitalization), reduce the likelihood to die.

It is interesting to note that the variable “CPAP time” has very sharp and narrow confidence intervals within the first five days (Figure 1). Oxygen is associated with increase in the likelihood to die, however this may be the result of interventions providing larger quantities of oxygen to the worst patients.

**Figure 1.**
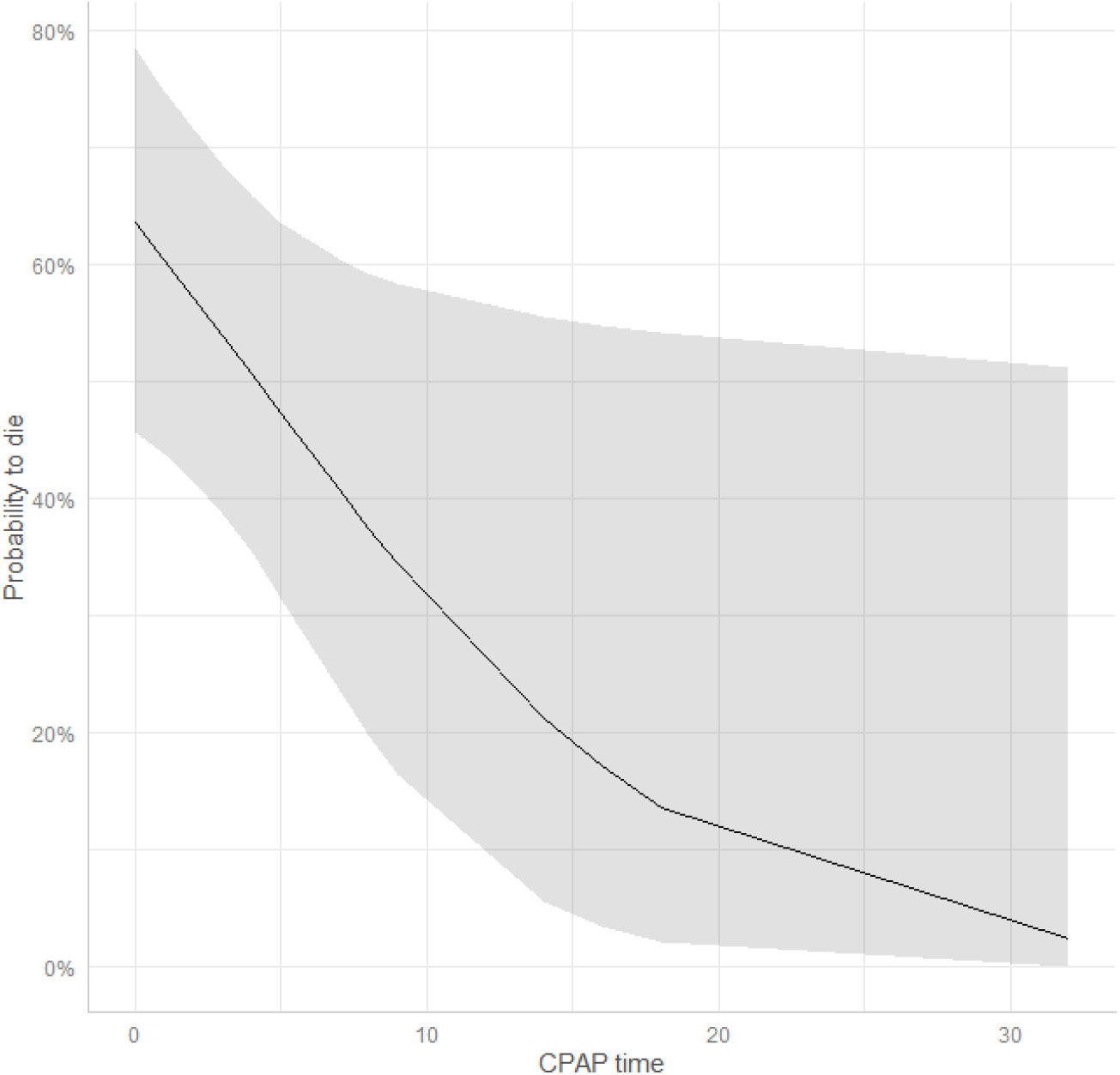
CPAP time effect on probability to die.

## Conclusion

There are no published trials of CPAP use in COVID-19 patients and as such the use of CPAP is based on clinical experience and “best endeavours”. Routine collection of data through the pandemic allowed for the comparison of changing clinical behaviours around the use of CPAP in COVID – 19.

The audit data presented indicates that there is clinical advantage in the early use of CPAP (defined as within 48 hours) in comparison to the late use of CPAP as indicated by comparing the magnitude of the odd ratios (Table 1).

Whilst this is a small data set and confounding factors are not included (drug administration and co-morbidities), although controlled by clinicians, it demonstrates evidence of clinical benefit and indicates a preference for early CPAP clinical strategy when treating patients with COVID-19. Further evidence for benefit should be sought from similar audits where data is available. A prospective trial of strategies would be appropriate in the future but will take time to design and approve. At this early stage in the treatment of COVID-19 respiratory infections the evidence presented supports the early use of CPAP in such patients.

## Data Availability

All data for the article is available from the authors.

## Acknowledgements

The outstanding contributions made by all members of our Multidisciplinary teams have been instrumental in introducing and developing this treatment intervention for our patients. Without their willingness to take this leap in faith adapting, learning and sharing their expertise, this improvement in care would not have been possible.

## Authors contribution

MF, RS and AA conceived of the study. JM contributed to refinement of the study protocol. AU collected and validated the data. LS analysed the data. LS and MF interpreted the statistical results. All the authors have edited and approved the final manuscript.

